# The endothelial growth factor Angiopoietin-2 is an accurate prognostic biomarker in patients with acetaminophen-induced acute liver failure

**DOI:** 10.1101/2025.02.19.25322567

**Authors:** David S. Umbaugh, Nga T. Nguyen, Steven C. Curry, Jody A. Rule, William M. Lee, Anup Ramachandran, Hartmut Jaeschke, Acute Liver Failure Study Group

**Affiliations:** Department of Pharmacology, Toxicology & Therapeutics, University of Kansas Medical Center, Kansas City, KS, USA; Department of Medical Toxicology, Banner – University Medical Center Phoenix, Phoenix, AZ, USA; Department of Medicine, and Division of Clinical Data Analytics and Decision Support, University of Arizona College of Medicine - Phoenix, Phoenix, AZ, USA; Division of Digestive and Liver Diseases, University of Texas Southwestern Medical Center, Dallas, TX, USA

**Keywords:** Drug hepatotoxicity, acetaminophen-induced liver injury, single cell RNA-sequencing, biomarkers

## Abstract

**Background and Aims:** Acetaminophen (APAP) overdose is the leading individual cause of acute liver failure (ALF) in the United States, with many patients rapidly progressing to hyperacute liver failure. While hepatocytes are the main target of APAP toxicity, endothelial cells (ECs) are also affected. However, the efficacy of an endothelial-specific biomarker to predict patient outcomes remains unknown. This study aimed to evaluate angiopoietin-2 (ANGPT2) as a prognostic biomarker for poor outcomes in APAP-induced ALF.

**Approach and Results:** Using human and mouse single-cell RNA sequencing (scRNAseq) data, we found that ANGPT2 expression was significantly elevated in ECs following APAP exposure. We measured circulating ANGPT2 levels from two independent APAP-ALF cohorts: a Phoenix cohort (n=43) and a cohort from the ALF Study Group (n=80). In the Phoenix cohort, ANGPT2 levels were significantly higher in non-survivors with an AUROC of 0.938. In the ALFSG cohort, we stratified patients based on time of symptom onset finding that ANGPT2 had improved prognostic value in early-presenting patients, with day 1 and day 3 AUC values of 0.825 and 0.918, respectively. Lastly, we combined the patient cohorts (n=110) finding that ANGPT2 alone or in combination with MELD outperformed MELD alone based on AUC (ANGPT2: 0.87, MELD 0.83, ANGPT2+MELD 0.90).

**Conclusions:** ANGPT2 is a promising prognostic biomarker for APAP-induced ALF, reflecting endothelial stress and offering superior predictive value compared to MELD alone, especially in early-presenting patients. Its capacity for predicting poor outcomes underscores its value in improving patient prognosis and therapeutic intervention strategies in APAP overdose cases.

**Lay Summary:** Accidental or intentional overdosing on acetaminophen can cause liver injury and in severe cases acute liver failure. Under these circumstances, receiving a liver transplant may be the only remaining therapeutic option. However, a liver transplant is a major surgery and commits the patient to a lifetime of anti-rejection medication. Because there is only a limited time window to decide who will recover and who needs a transplant to survive, prognostic biomarkers are essential to identify transplant candidates as early as possible after the overdose. In this study we discovered that plasma levels of the endothelial growth factor angiopoietin-2 can accurately predict at the peak of injury who will need a liver transplant to survive. In addition, this biomarker can be rapidly measured, which allows the data to be available for clinical decision making.

**Highlights:**

- Acetaminophen-induced liver injury can cause hyper-acute liver failure within 3 to 7 days with a high probability of negative outcome.
- Under these conditions, a liver transplant may be the only therapeutic option.
- In two independent cohorts, angiopoietin 2 was identified as an early prognostic biomarker for poor outcome.
- Angiopoietin can more accurately inform clinical management during the initial stages of hospital presentation than the MELD score.

## INTRODUCTION

Acute liver failure is characterized by extensive hepatocyte death causing coagulopathy and hepatic encephalopathy without preexisting liver disease (1). Though the primary individual cause of ALF in the United States is an acetaminophen (N-acetyl-p-aminophenol/APAP) overdose, only 16% of all liver transplant patients have an etiology of an APAP overdose (2).

This is frequently due to the rapid progression of sequelae after an APAP-induced liver injury. While ALF is defined as liver failure in less than 28 days, APAP overdose patients tend to experience a much more rapid onset of liver failure occurring in 3-7 days, termed hyperacute liver failure, which is driven by the massive centrilobular necrosis that is a key characteristic feature of an APAP overdose (1). Because of this, clinical decisions regarding the utility of liver transplantation in these patients must be made quickly and in the absence of significant longitudinal data. This decision-making process is further confounded by the fact that most APAP overdose patients will survive if they receive the standard of care, N-acetylcysteine (NAC) (3,4) and ICU support. Besides the initial dose consumption of APAP and time to presentation to the clinic, other factors helping differentiate which APAP overdose patients survive while others progress to multi-organ failure and death are unclear. As such, there remains an urgent clinical need to identify prognostic biomarkers which can distinguish patients with pathological progression towards poor outcome.

Although hepatocytes are the prototypical target of APAP toxicity, sinusoidal endothelial cells (ECs) also experience APAP-induced toxicity. Microvascular injury occurs prior to hepatocyte injury (5) and involves the formation of peroxynitrite (6), a key molecular event in the APAP toxicity cascade (7,8). The swelling of ECs, loss of fenestrae and decreased blow flow occur in the first 2-3 hours following APAP exposure (9,10), with greater impairment in the pericentral endothelial cells (PC-ECs) (10). While several mechanistic biomarkers have been established for different facets of APAP toxicity, such as mtDNA (11) or carbamoyl phosphate synthetase 1 (12) for mitochondrial dysfunction, high mobility group box protein 1 (13) or nuclear DNA (14) for hepatocyte necrosis, or α-fetoprotein or osteopontin (15) for hepatocyte proliferation, no similarly defined EC related biomarker has been defined for APAP.

Given that proxy measures of hepatocyte injury, such as ALT, have limited prognostic value for determining patient outcome (14,16,17), the extent of liver injury is not the only factor influencing development of ALF. This suggests the involvement of additional factors in the initiation or exacerbation of biological events in the subset of patients who progress to poor outcome. In this vein, we have recently identified the novel prognostic biomarker, CXCL14 (17), which we demonstrated was related to the initiation of a p21-dependent secretome in the subset of APAP-ALF patients with poor outcome (18). Correspondingly, the question emerged whether this strata of APAP-ALF patients experience greater EC stress and if it could be represented by a circulating factor.

Angiopoietin-2 (ANGPT2) is an endothelial specific factor sitting at the nexus of angiogenesis and inflammation (19). It is stored in Weibel-Palade bodies of ECs and is rapidly released in response to stimulation (20). ANGPT2 promotes cell adhesion by sensitizing ECs to TNFα, playing a critical role in regulating the inflammatory response (21). Elevated levels of circulating ANGPT2 are associated with liver angiogenesis and fibrosis in chronic hepatitis C patients (22). We evaluated both mouse and human APAP overdose single-cell RNA sequencing (scRNAseq) data finding that *Angpt2* is highly expressed in ECs, specifically PC-ECs, and that PC-ECs exhibit sustained stress following APAP exposure. Taken together, this led us to hypothesize that ANGPT2 may be an effective prognostic biomarker, as it may be reflective of an exacerbated EC stress in the subset of APAP OD patients with poor outcomes. Our evaluation of circulating ANGPT2 in two separate APAP-induced ALF cohorts revealed that ANGPT2 is an effective prognostic biomarker, outperforming MELD in this large patient cohort.

## METHODS

### Patient information

The patient samples and data in this study were obtained from two different cohorts: (A) plasma samples obtained from 43 APAP overdose patients at the Banner - University Medical Center Phoenix in Phoenix AZ, USA, and (B) 80 APAP overdose patient serum samples obtained through the ALF Study Group (ALFSG) registry. Summary information for the Phoenix Cohort is described in Table 1 and summary information for the ALFSG Cohort in Table 2. In the Phoenix Cohort, diagnosis for APAP overdose was made by a physician based on reported history of APAP overdose, detectable plasma APAP levels, and/or peak ALT level ≥ 1000 IU/L. Heparinized collection tubes were used to collect blood samples at study admission and every 24 h after until patient death or discharge. The whole blood was centrifuged for 10 min at 1,000g to obtain plasma. In the ALFSG Cohort, site investigators used standard criteria for diagnosis of APAP overdose, which included a history of APAP overdose, detectable APAP levels in circulation, and/or an ALT level ≥ 1,500 IU/L. Inclusion criteria for enrollment into the registry included: severe liver injury, INR ≥ 1.5, hepatic encephalopathy, and the onset of liver failure within 26 weeks of illness without chronic liver disease. Written consent was obtained from next of kin due to impaired capacity of the patients due to APAP-induced ALF. The patient samples and data for both cohorts were acquired with informed consent and approved by Institutional Review Boards in adherence to the 1975 Declaration of Helsinki.

**Table 1.** Patient and clinical characteristics of the Phoenix Cohort.

|  |  | Survivors | Nonsurvivors | p |
| --- | --- | --- | --- | --- |
| Male, n |  | 15 | 4 | 0.116 |
| Female, n |  | 13 | 11 |  |
| APAP peak level (mg/L) |  | 28 (72) | 51 (132) | 0.049 |
| Weight (kg) |  | 79 (21) | 72 (19) | 0.03 |
| BMI |  | 27 (8) | 26 (2) | 0.30 |
| ALT<br>(IU/L) | Day 1 | 6904 (3875) | 4732 (3469) | 0.27 |
|  | Day 2 | 4602 (2873) | 3979 (4121) | 0.54 |
|  | Day 3 | 3219 (1632) | 2257 (2107) | 0.44 |
| AST<br>(IU/L) | Day 1 | 6909 (7847) | 7927 (11218) | 0.39 |
|  | Day 2 | 2610 (5058) | 6896 (10368) | 0.05 |
|  | Day 3 | 587.5 (1626) | 2350 (3606) | 0.04 |
| MELD | Day 1 | 23 (19) | 38 (10) | 0.02 |
|  | Day 2 | 24 (16) | 43 (17) | 0.0009 |
|  | Day 3 | 19 (15) | 44 (11.5) | 0.0004 |
| INR | Day 1 | 2.7 (3.85) | 6.4 (3.75) | 0.19 |
|  | Day 2 | 2.3 (2.4) | 6.8 (11.8) | 0.03 |
|  | Day 3 | 1.95 (1.35) | 4.7 (3.9) | 0.002 |
| Bilirubin<br>(mg/dL) | Day 1 | 2.6 | 4.8 | 0.03 |
|  | Day 2 | 2.8 | 5.2 | 0.07 |
|  | Day 3 | 3.7 | 6.4 | 0.32 |

Data is expressed as median with the difference from the third and first quartile (Q3-Q1) for continuous variables. The p value is calculated by Fisher exact test for categorical variables and by the Wilcoxon rank-sum test for continuous variables (Survivors vs Non-survivors).

**Table 2.** Patient and clinical characteristics of the ALFSG Cohort.

|  |  | Survivors | Nonsurvivors (or LT) | p |
| --- | --- | --- | --- | --- |
| Male, n |  | 15 | 9 | 0.33 |
| Female, n |  | 27 | 29 |  |
| APAP peak level (mg/L) |  | 18.35 (37.5) | 22.5 (52) | 0.85 |
| Weight (kg) |  | 73 (29.5) | 67.4 (18) | 0.02 |
| BMI |  | 28 (9.0) | 25 (6.1) | 0.03 |
| ALT (IU/L) | Day 1 | 3635 (4010) | 4010 (2747) | 0.38 |
|  | Day 3 | 1512 (1685) | 1832 (1344) | 0.30 |
| AST (IU/L) | Day 1 | 2600 (3451) | 4799 (5615) | 0.007 |
|  | Day 3 | 347 (683) | 1151 (1567) | 0.007 |
| MELD | Day 1 | 34 (21) | 44 (10) | 0.0006 |
|  | Day 3 | 32 (26) | 44 (9) | 0.0003 |
| Creatinine (mg/dL) | Day 1 | 1.23 (1.38) | 1.86 (1.8) | 0.14 |
|  | Day 3 | 0.99 (1.15) | 1.39 (1.7) | 0.23 |
| Bilirubin (mg/dL) | Day 1 | 4.8 (3.9) | 5 (4.4) | 0.23 |
|  | Day 3 | 4.7 (5.3) | 8.9 (6.2) | 0.00096 |

Data is expressed as median with the difference from the third and first quartile (Q3-Q1) for continuous variables. The p value is calculated by Fisher exact test for categorical variables and by the Wilcoxon rank-sum test for continuous variables (Survivors vs Non-survivors).

### Animals

Eight-week-old male C57BL/6 J mice (Jackson Laboratories, Bar Harbor, Maine) were kept in an environmentally controlled room with a 12 h light/dark cycle and *ad libitum* access to food and water. All experimental protocols were approved by the Institutional Animal Care and Use Committee of the University of Kansas Medical Center and followed the criteria of the National Research Council for the care and use of laboratory animals.. Mice were intraperitoneally (i.p.) injected with 300 mg/kg APAP (Sigma-Aldrich, St. Louis, MO) or saline vehicle after overnight fasting.

### Histology and immunohistochemistry

Mouse liver tissue was formalin-fixed and embedded in paraffin and sectioned at 5 µm. For staining, liver sections were hydrated, and antigen retrieval was performed by boiling in sodium citrate buffer (10 mM sodium citrate, 0.05% Tween 20, pH 6.0). The liver sections were treated with 3% hydrogen peroxide, followed by TBS washes, and blocked in 3% BSA and 5% goat serum for 1h at room temperature. Sections were incubated with rabbit anti-CD31 (Cell Signaling Technologies, #77699, 1:200) overnight at 4° Celsius. The next day SignalStain Boost detection reagent (Cell Signaling Technologies, #8114) was added for 30 min, followed by visualization using SignalStain Dab Chromogen concentrate (Cell Signaling Technologies, # 8059) with images captured using an Olympus BX51TF.

### ELISA assay

Circulating ANGPT2 was measured by ELISA using an R&D ANGPT2 assay (#DY623) for the serum and plasma samples. The protocol was followed according to the manufacturer’s instructions. In brief, samples were pipetted in a 96-well plate using a solid phase sandwich ELISA and then sealed for overnight incubation. The next day the plate was washed three times with PBS followed by blocking (1% BSA) at room temperature for 1 hour. Plasma or serum (100 µL) was added to plate in duplicate after dilution (dilution was optimized either 1:5, 1:10 or no dilution in 1% BSA) and incubated at room temperature for 2 hours. The plate was washed, and detection antibody was added for 2 hours. Streptavidin-HRP was added to increase the signal intensity and tetramethylbenzidine substrate was added to initiate the reaction for a 20-minute incubation at room temperature. All standards, samples, and controls were run in duplicate and then averaged and subtracted from the zero standard optical density. A four-parameter logistic curve was constructed as the standard to derive ANGPT2 concentration.

### Data analysis of Phoenix and ALFSG cohorts

Area under the receiver-operating characteristics curve analysis (AUROC) and classification performance metrics were determined in R version 4.1.0 using the R packages caret (21) and pROC (24). For the analysis of the combined cohort (n=110) and subset cohort (n=63), data preprocessing included scaling, centering, and imputation by bagging of regression trees using Caret’s *preProcess* function. The logistic regression models were evaluated based on: AUROC, sensitivity, specificity, and accuracy. The 95% confidence interval of the AUROC was determined by bootstrapping (2000 replicates). The selected threshold was based on maximizing the distance to the identity line on the ROC curve (Youden’s J statistic).

### Human single-cell sequencing datasets

Human single cell data for Supplemental Figure 1 (B-D) was obtained as processed, annotated datasets from the Chan Zuckerberg CELL by GENE Discover initiative. The single-cell data for Supplemental Figure 1A and E was obtained directly from GEO at GSE223560. Seurat’s *AggregateExpression* function was used on each individual dataset to create pseudo-bulk gene expression profiles for each individual sample. The sample size for each group is described in the figure legends. For Supplemental Figure 2, data was obtained from GSE223560 consisting of human single-nuclei RNAseq (snRNAseq) from healthy (n=9) and APAP-ALF (n=11) subjects. The provided cell labels were used to extract the ECs.

The ECs were re-processed following standard Seurat processing (25) with integration using Harmony (26). Pathways relating to angiogenesis and inflammation were acquired through the Molecular Signatures Database (MsigDB) for input for gene set variation analysis (GSVA).

Significant gene sets were then used to calculate the Angiogenesis or Inflammatory Response score using Seurats *AddModuleScore* function (27). All statistical analysis and figure production was performed in R (v 4.1.0).

### Mouse APAP single-cell RNA-sequencing Atlas

The mouse single cell RNAseq atlas consists of three integrated datasets GSE136679, zenodo.6035873, and GSE255835 with data pre-processing, integration, and cell annotation previously described (18). The ECs were extracted from the scRNAseq mouse APAP atlas consisting of 4704 control ECs and 5865 APAP ECs.

Endothelial cells were classified into 5 groups based on their spatial characteristics: arterial ECs, periportal ECs (PP-ECs), midzonal ECs (MZ-ECs), pericentral ECs (PC-ECs), pericentral venous ECs (CV-EC) and a separate group identified as cycling ECs. *Vwf* readily identified arterial ECs, *Ntn4*, *Msr1*, and *Efnb2* identified PP-ECs, *Stab2* and *Lyve1* identified MZ-ECs, *Wnt2*, *Kit* and *Thbd* identified PC-ECs, CV-ECs by *Wnt9b* and *Rspo3*, and *Mki67*, *Pclaf* identified cycling ECs. ECs were labeled based on marker gene expression (28) using Seurat’s *AddModuleScore* function. All statistical analysis and figure production was performed in R (v 4.1.0).

### Threshold analysis

We used the cutpointr package (version 1.1.2) in R version 4.1.0 to determine the threshold values for ANGPT2. The optimal threshold value was based on maximizing the sum of sensitivity and specificity or maximizing accuracy. The cutpointr function was run with stratified bootstrapping (1000) to ensure a similar proportion of both positive and negative cases in each resample.

### Statistics

Continuous variables are reported as median with the difference from the third and first quartile (Q3-Q1) and evaluated by Wilcoxon-rank sum test. The p value for categorical variables was determined using Fisher’s exact test. Multiple group comparisons were evaluated by two-way ANOVA with post hoc testing (Tukey or Sidak). The logistic regression models were evaluated using AUROC and other metrics relating to classification. A two-tailed alpha was set at 0.05. Statistical analysis was performed in GraphPad Prism 10 or using R version 4.1.0.

## RESULTS

### ANGPT2 is expressed in pericentral endothelial cells and increases after APAP

Our evaluation of three independent human scRNAseq datasets revealed that *Angpt2* is expressed highly in both endothelial cells (ECs) and hepatocytes (Supp Fig 1A-C). Previous reports regarding the role of endothelial zonation in liver diseases (28) led us to investigate if *Angpt2* expression differed in ECs based on zonal location. scRNAseq analysis of twenty-four human livers demonstrated that *Angpt2* expression is higher specifically in pericentral (PC) endothelial cells (PC-ECs) (Supp Fig 1D). In human APAP livers, the highest expression of *Angpt2* was in ECs compared to other liver cell types, including hepatocytes (Supp Fig 1E). Comparative analysis of ECs from human single nuclei RNA seq (snRNAseq) of APAP explant livers revealed several distinct clusters (Supp Fig 2A-C), enriched in angiogenesis and inflammatory processes (Supp Fig 2D-G). Since APAP pathophysiology in the mouse model closely replicates changes in humans (29), we turned to our scRNAseq APAP hepatotoxicity mouse atlas, consisting of cells from 24 h to 96 h after APAP, to evaluate the temporal dynamics of *Angpt2* after an APAP overdose. We separated the EC population and defined spatial populations based on marker genes (Fig 1A-B). We found that *Angpt2* expression increased in both PC-EC and periportal ECs (PP-ECs) at 24 h after APAP, before returning to baseline levels by 96 h after APAP (Fig 1C). Moreover, cell abundance analysis revealed a decrease in the proportion of PC-EC at 24 h, which we confirmed by staining for the pan-EC marker CD31 (Fig 1D-E). A global analysis comparing PC-ECs and PP-ECs at each timepoint revealed an initial stress in both PC-ECs and PP-ECs at 24 h, with a persistent stress signature in PC-ECs (Fig 1F). Moreover, angiogenic signaling was induced in both PC and PP-ECs at 24 h with a more robust induction in PC-ECs persisting until 96 h (Fig 1G). Collectively, this suggests that PC-ECs are uniquely susceptible to APAP-induced stress, highlighting that secreted angiogenic factors such as ANGPT2 may be a prognostic biomarker in APAP-induced ALF.

**Figure 1:**
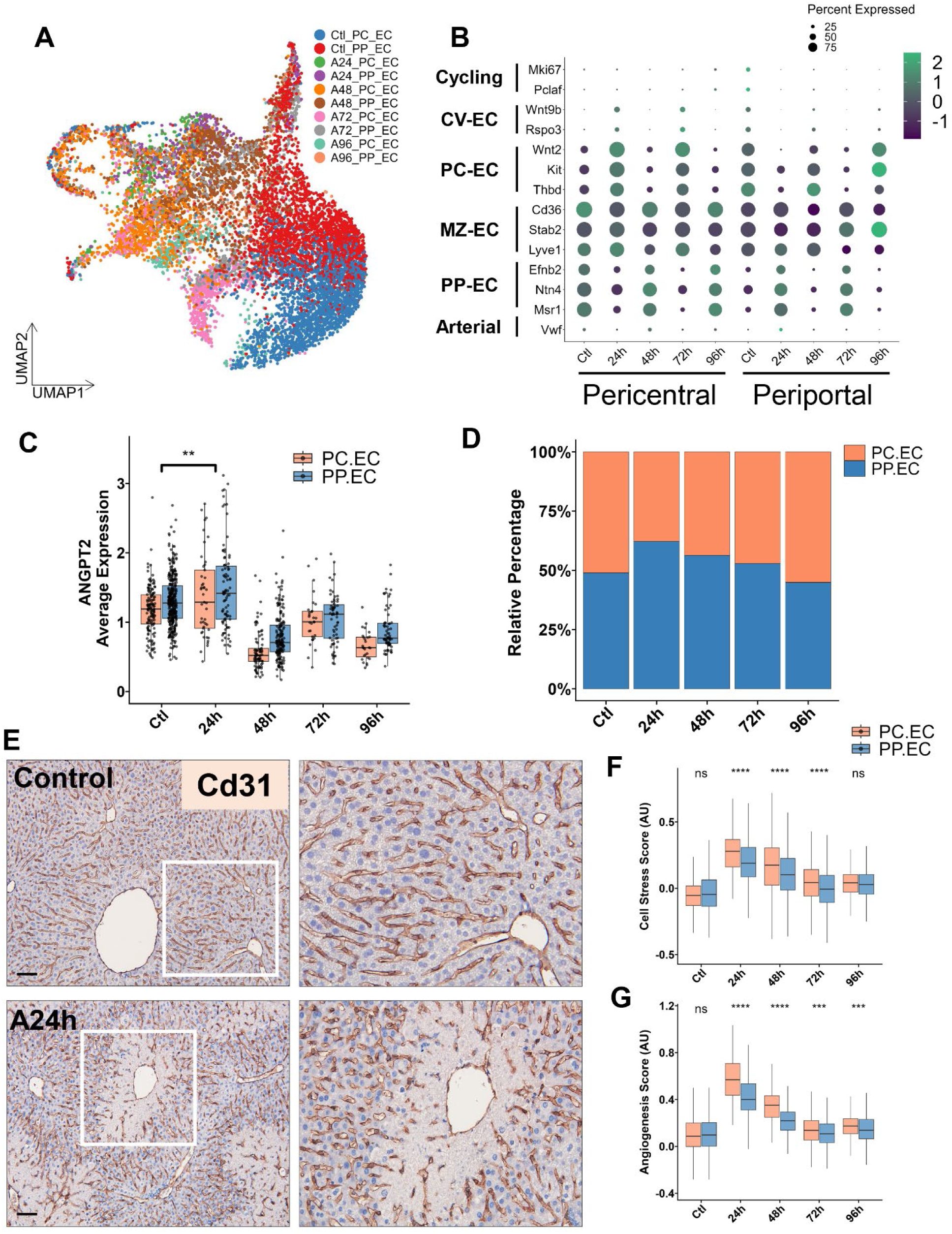
Spatial mapping of mouse endothelial cells reveals sustained induction of angiogenesis and stress in pericentral ECs. (A) UMAP of ECs across the APAP toxicity time course (24h to 96h) labeled by lobular location. (B) Dotplot showing marker genes for different EC subpopulations. (C) Boxplot for the average expression of *Angpt2* across time and lobular location. (D) Proportion bar chart showing the change in relative abundance of PP and PC ECs. (E) Representative IHC staining for the pan-EC marker CD31 (left: 200x, scale bar = 100 µm, right: 400x). (F-G) Boxplot for cell stress (F) or angiogenesis (G) calculated using the *AddModuleScore* function from Seurat. Statistical significance was assessed using the non-parametric Wilcoxon rank sum test comparing control vs 24h (C) or in (F-G) comparing PC-EC vs PP-EC at the respective timepoint.

**Figure 2:**
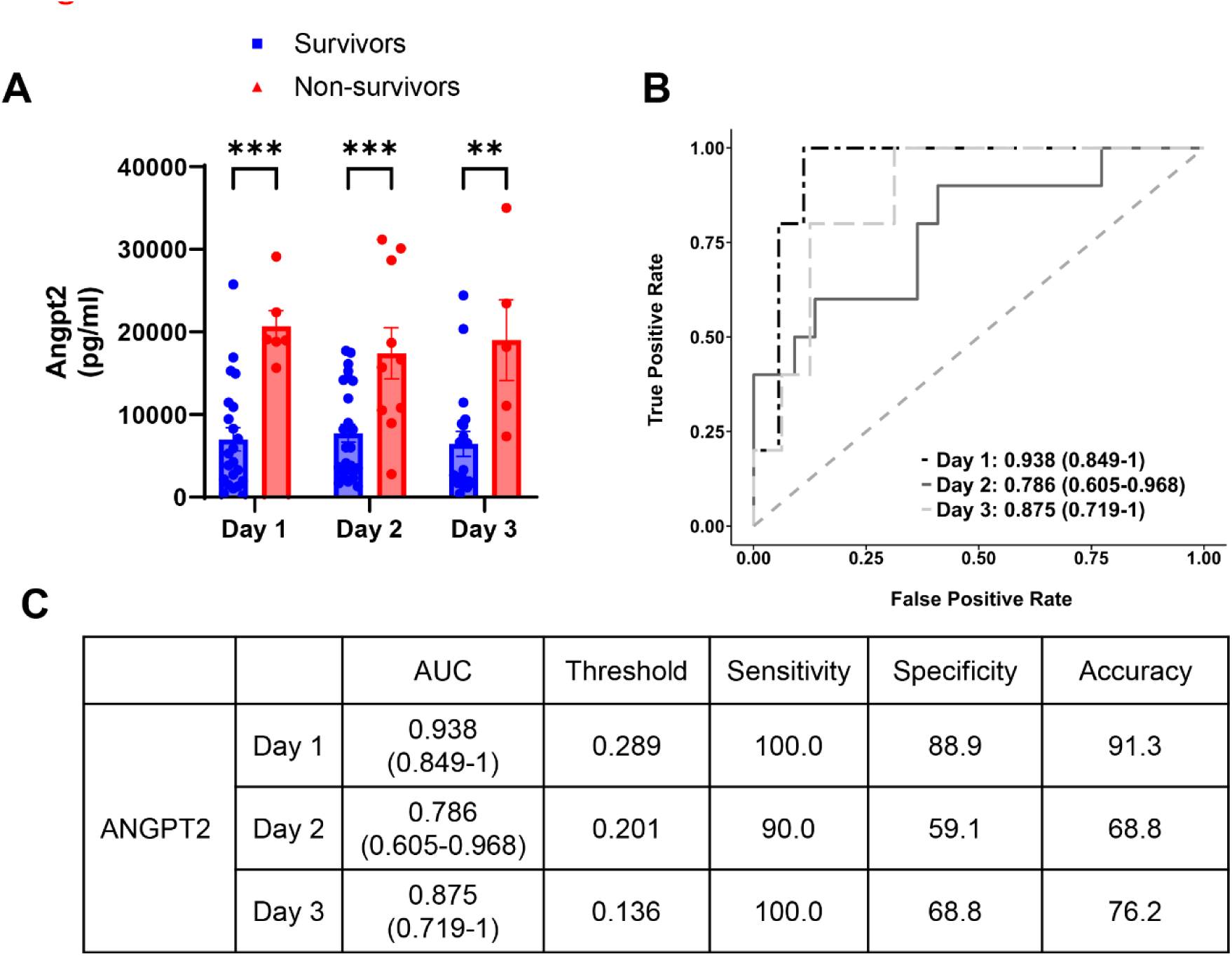
Evaluation of circulating ANGPT2 between APAP overdose survivors and non-survivors in Phoenix Cohort. (A) Plasma ANGPT2 assessed by ELISA. (B) AUROC based on ANGPT2 (AUC is shown with 95% CI based on 2000 stratified bootstrap replicates). (C) Data table summarizing AUC, sensitivity, specificity, and accuracy at thresholds that maximized the Youden’s J statistic. Data are presented as mean ± SEM, *p<0.05, **p<0.01, ***p<0.001.

### ANGPT2 differentiates APAP overdose patients with poor outcome from survivors

The Phoenix Cohort consists of 28 surviving APAP OD patients and 15 patients with poor outcome (non-survivors). In this patient cohort, the patients were immediately enrolled for the study as APAP OD patients after arriving at the hospital. Clinical measurements were taken every 24 hours after admission. The measurements and demographic information are reported in Table 1. We found plasma ANGPT2 was higher at day 1, 2, and 3 in non-survivors compared to the survivors (Fig 2A). ROC analysis of ANGPT2 was highest at day 1 and day 3 (Day 1 AUC: 0.938, Day 3 AUC: 0.875), demonstrating its utility as a prognostic biomarker in this patient cohort (Fig 2B-C).

### Confirmation of ANGPT2 as a prognostic biomarker in a larger ALFSG Cohort

To validate our findings, we assessed 80 patient samples obtained through the ALFSG Registry which consisted of 42 survivors and 38 patients with poor outcome (non-survivors or liver transplant). The demographic and clinical measurements are presented in Table 2. We evaluated ANPGT2 serum levels in the ALFSG Cohort on day 1 and day 3 (after enrollment), finding that ANGPT2 was increased in patients with poor outcome (Fig 3A). Moreover, at both day 1 and 3 ROC analysis yielded a similar AUC of 0.748 and 0.757, respectively (Fig 3B-C). However, we and others have previously found that the prognostic value for these biomarkers can be influenced based on the timing of the APAP overdose relative to presentation in the clinic (17, 30), likely due to the differences in biological events which occur at different phases of APAP toxicity (31). As such, we stratified the ALFSG Cohort into two groups based on reported onset of symptoms, either less than or equal to two days (n=31) or more than two days (n=48). In this subset ROC analysis, we found in the early presenting patients that ANGPT2 had improved performance for distinguishing patients with poor outcome (Day 1 AUC: 0.825, Day 3 AUC: 0.918) (Supp Fig 3A-B) with a corresponding decrease in relative performance in late presenting patients (Day 1 AUC: 0.707, Day 3 AUC: 0.656). To explore this further, we also performed a sub-analysis based on reported timing of the APAP overdose, stratifying patients as early (≤ 3 days post APAP OD) or late (>3 days). Again, ANGPT2 performed better in early patients compared to late (Supp Fig 3 C-D, Day 1 AUC: 0.739 vs 0.655, Day 3 AUC: 0.814 vs 0.50).

**Figure 3:**
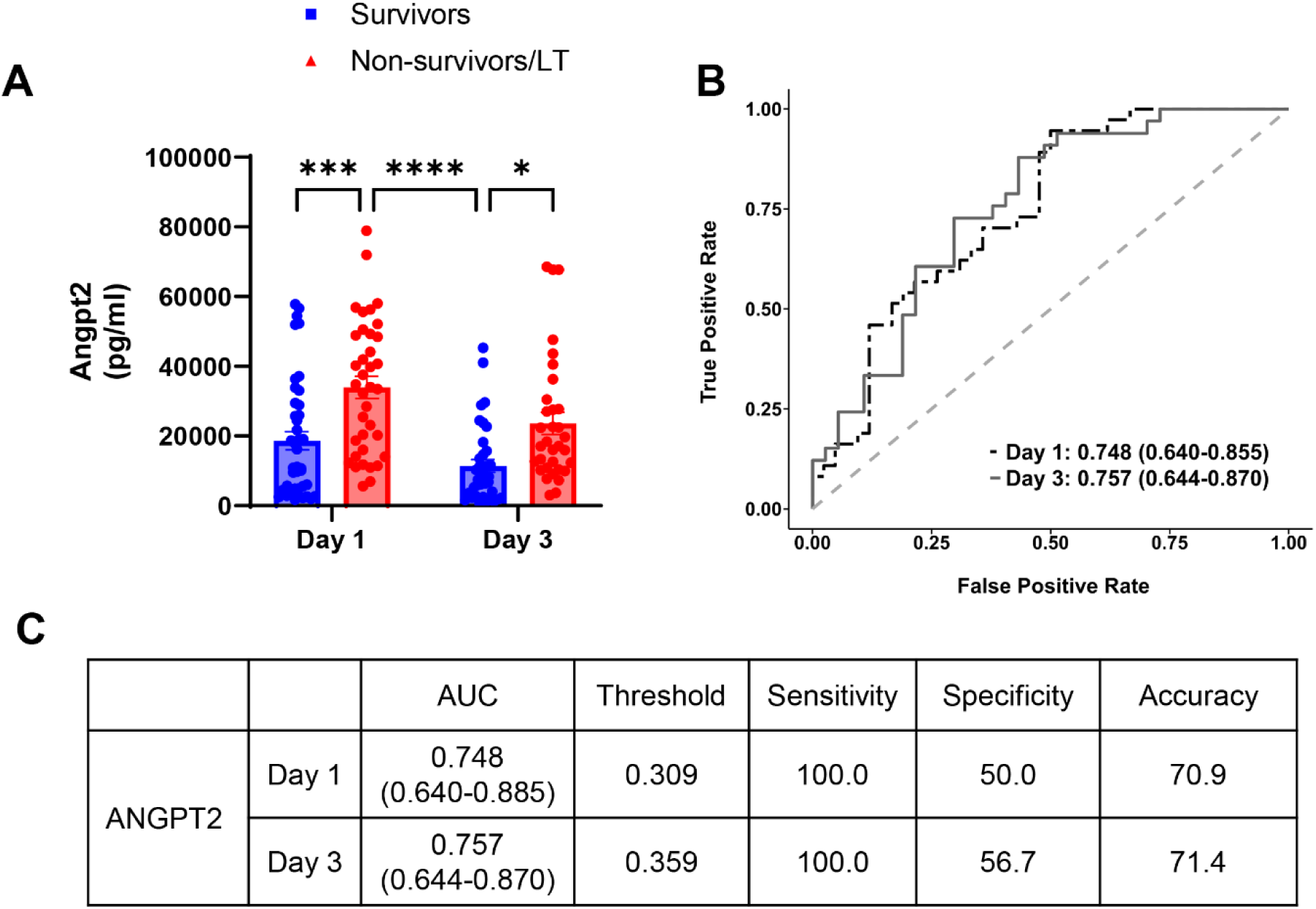
Circulating ANGPT2 distinguishes survivors and non-survivors/LT in the ALFSG Cohort. (A) Serum ANGPT2 assessed by ELISA. (B) AUROC based on ANGPT2 (AUC is shown with 95% CI based on 2000 stratified bootstrap replicates). (C) Data table summarizing AUC, sensitivity, specificity, and accuracy at thresholds that maximized the Youden’s J statistic. Data are presented as mean ± SEM, *p<0.05, **p<0.01, ***p<0.001.

Notably, using either symptom onset or APAP OD to identify early presenting patients (Table 3 and 4), ANGPT2 outperformed MELD in the early patient cohort subset (Symptom Onset: ANGPT2 vs MELD Day 1: 0.825 vs 0.764, Day 3: 0.918 vs 0.757, APAP OD: Day 1: 0.739 vs 0.73, Day 3: 0.814 vs 0.705).

**Table 3.** AUROC analysis in a subset of patients in the ALFSG cohort with reported onset of symptoms.

|  |  | Early | Late |
| --- | --- | --- | --- |
| ANGPT2 | Day 1 | 0.825 (0.676-0.974) | 0.707 (0.558-0.856) |
|  | Day 3 | 0.918 (0.8 – 1.0) | 0.656 (0.491-0.821) |
| MELD | Day 1 | 0.764 (0.578-0.95) | 0.746 (0.597-0.895) |
|  | Day 3 | 0.757 (0.573-0.941) | 0.749 (0.597-0.901) |

Early patients presented to the clinic within two days of symptom onset (n=31), while late patients presented after more than two days (n=48). AUC is shown with the 95% CI in parentheses.

**Table 4.**
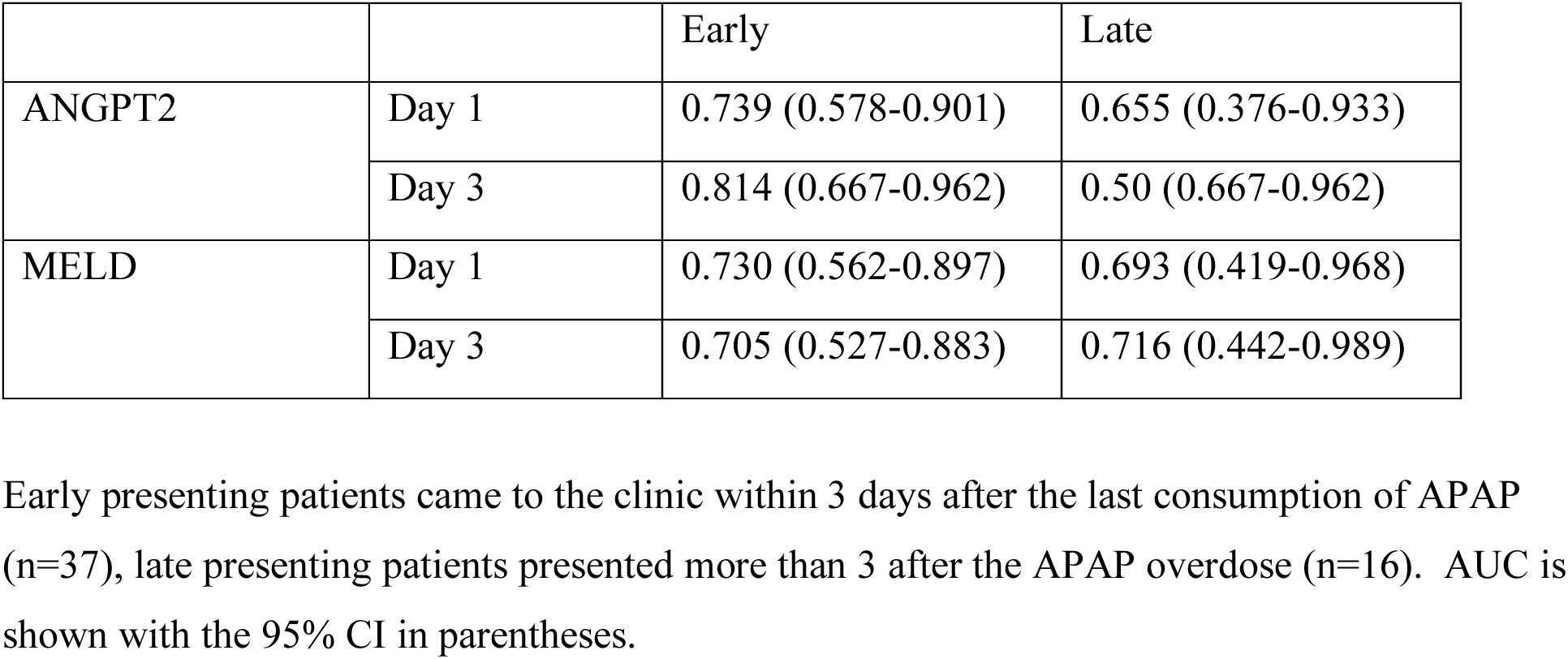
AUROC analysis from a subset of patients in the ALFSG cohort with reported timing of APAP overdose.

|  |  | Early | Late |
| --- | --- | --- | --- |
| ANGPT2 | Day 1 | 0.739 (0.578-0.901) | 0.655 (0.376-0.933) |
|  | Day 3 | 0.814 (0.667-0.962) | 0.50 (0.667-0.962) |
| MELD | Day 1 | 0.730 (0.562-0.897) | 0.693 (0.419-0.968) |
|  | Day 3 | 0.705 (0.527-0.883) | 0.716 (0.442-0.989) |

Early presenting patients came to the clinic within 3 days after the last consumption of APAP (n=37), late presenting patients presented more than 3 after the APAP overdose (n=16). AUC is shown with the 95% CI in parentheses.

### ANGPT2 outperforms MELD as a prognostic biomarker in the combined cohort

To synthesize our findings, we performed ROC analysis on three different logistic regression models using either ANGPT2, MELD, or ANGPT2 and MELD in the combined Phoenix and ALFSG Cohort (n = 110). The performance metrics for all three models were comparable in terms of AUC and accuracy at both day 1 and day 3 measurements with the combination model (ANGPT2+MELD) yielding the highest AUC at day 3 (ANGPT2+MELD: 0.83, ANGPT2: 0.77, MELD: 0.79) (Fig 4A-C). Next, we took the same subset of ALFSG patients (reported onset of symptoms less than 3 days) in combination with the Phoenix Cohort (n = 63) and performed the ROC analysis across the three different models. We found that ANGPT2 alone or in combination with MELD outperformed MELD alone at both day 1 and day 3 when comparing AUC (Day 3 ANGPT2: 0.87, MELD: 0.83, ANGPT2+MELD: 0.90) (Fig 5A-C). Collectively, these findings highlight that ANGPT2 is comparable to MELD when the analysis is timing agnostic, however, ANGPT2 or in combination with MELD is superior to MELD alone when stratifying patients based on additional clinical information.

**Figure 4:**
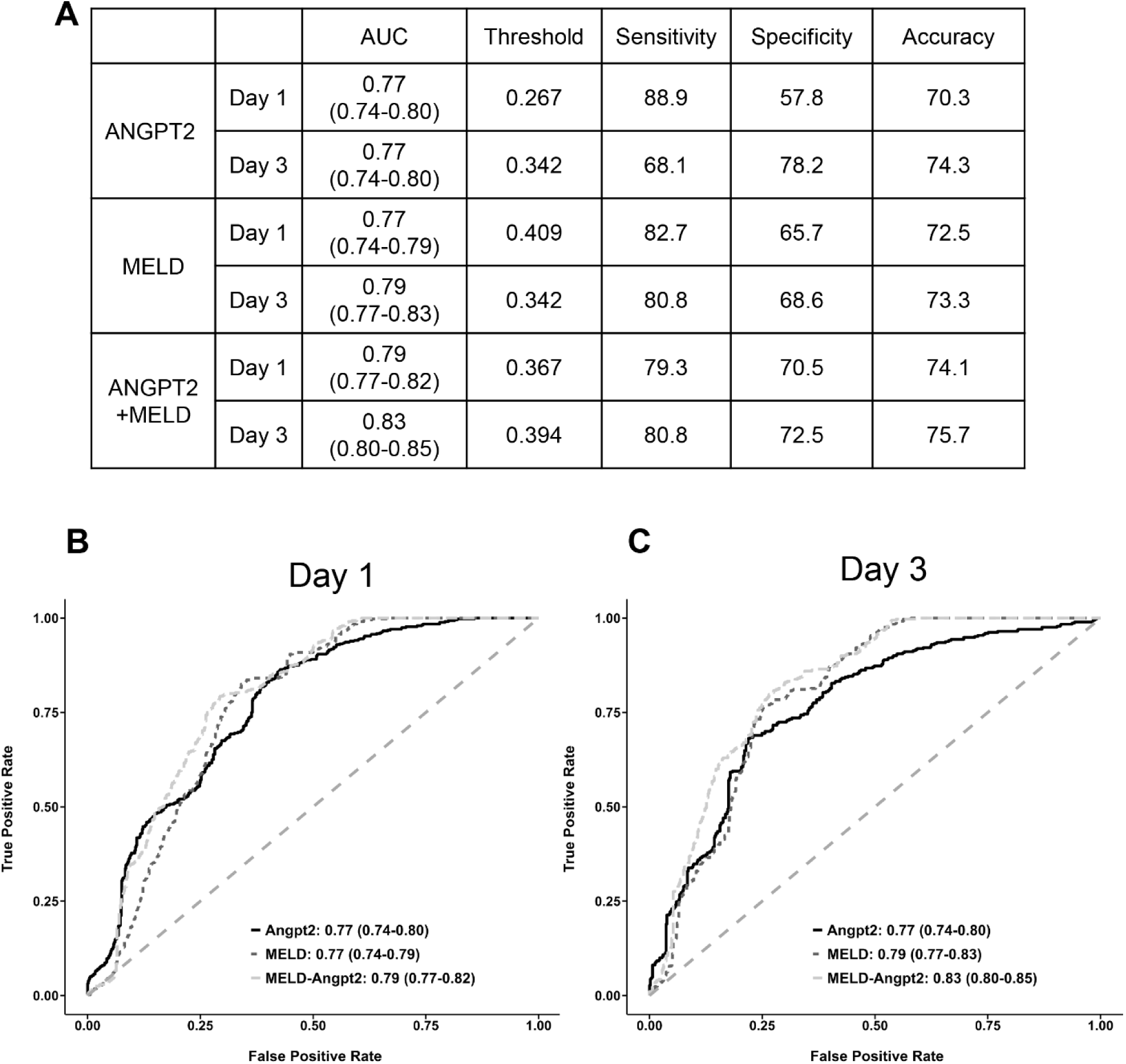
Combination model of ANGPT2 and MELD accurately predicts patient outcome in combined patient cohorts. (A) Data table summarizing AUC, sensitivity, specificity, and accuracy at thresholds that maximized the Youden’s J statistic (n=110 on Day 1, n=94 on Day 3). AUROC for each logistic regression model on Day 1 (B) or (C) Day 3.

**Figure 5:**
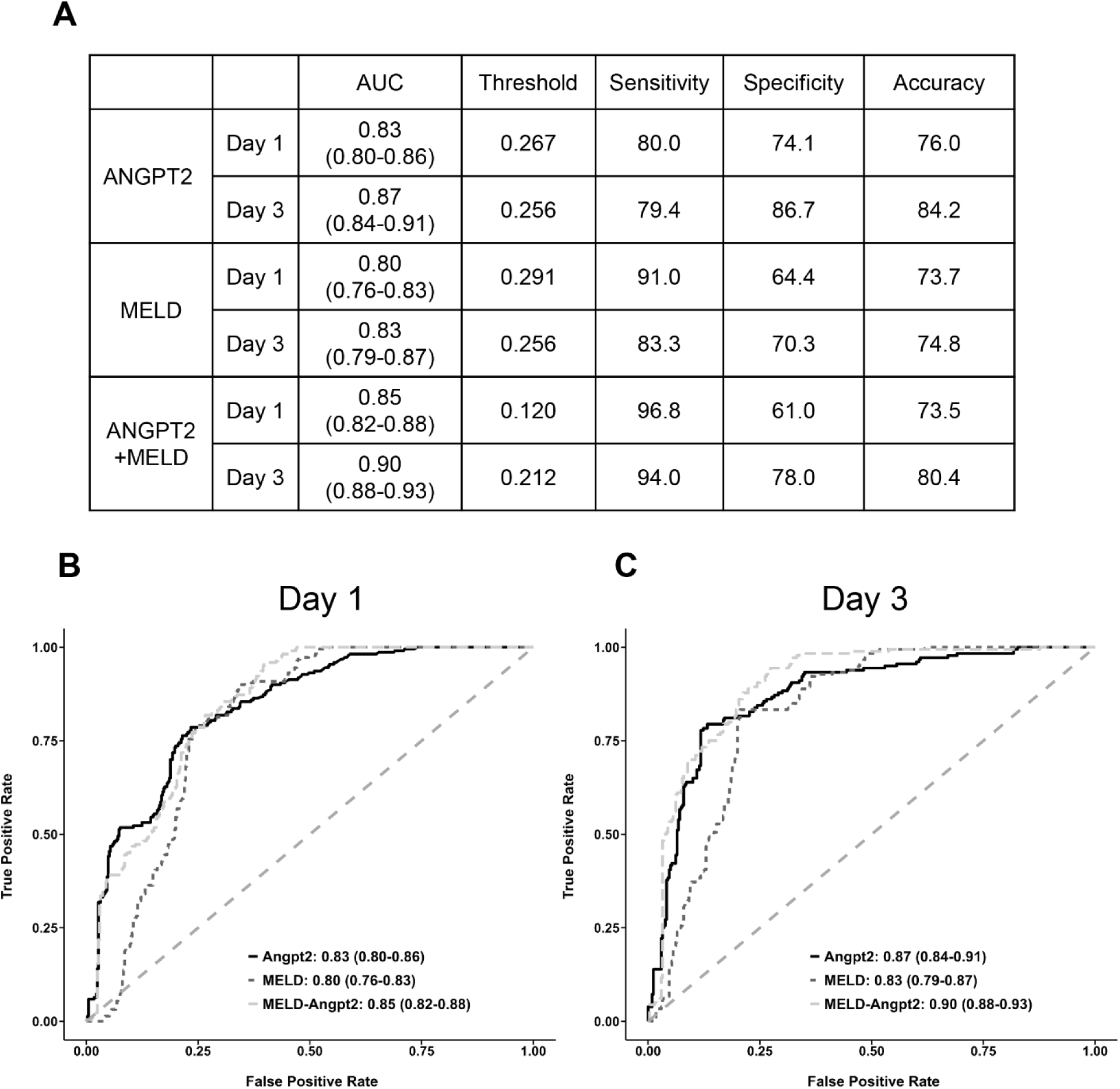
ANGPT2 outperforms MELD in the subset cohort with early presenting patients. (A) Data table summarizing AUC, sensitivity, specificity, and accuracy at thresholds that maximized the Youden’s J statistic (n=63 on Day 1, n=52 on Day 3). AUROC for each logistic regression model on Day 1 (B) or Day 3 (C).

### Defining ANGPT2 threshold values

Next, we sought to establish threshold values for ANGPT2 which could be used to distinguish patients with poor outcome. The commonly used cutoff values for MELD are ≥ 20 or ≥ 30, with the former often resulting in higher sensitivity but lower specificity, while the latter the inverse (15). We evaluated threshold values for ANGPT2 in both the entire combined cohort (n=110) (Fig 6) and in the subset cohort (n=63) at both day 1 and day 3 (Fig 7). The optimal threshold value for ANGPT2 was remarkably stable, around 11000 pg/ml at both day 1 in the full cohort (Fig 6A-C), and day 1 and 3 in the subset cohort (Fig 7A-F) (Full Cohort Day 1: 10975 pg/ml, Subset Cohort, Day 1: 11112 pg/ml, Day 3: 10904 pg/ml). On day 3 in the full cohort the optimal threshold value was slightly lower, ANGPT2 ≥ 9988 (Fig 6 D-F). For comparison, the performance of the MELD cutoff is reported (Fig 6 C, F and Fig 7 C, F). Notably, ANGPT2 has a higher overall accuracy across all conditions. Taken together, we establish a threshold value of ANGPT2 to help guide clinical management, though future validation in larger cohorts may be warranted.

**Figure 6:**
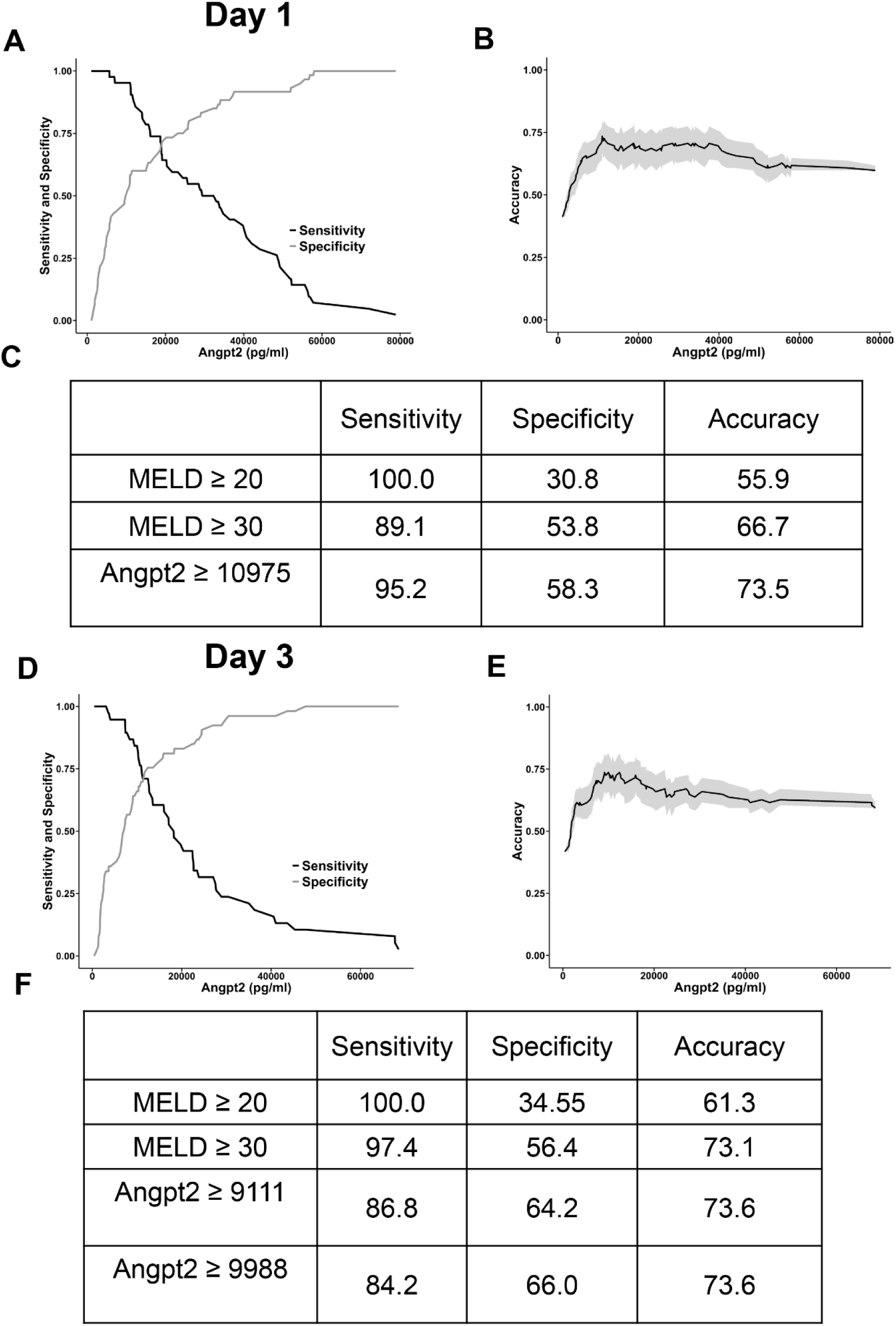
Performance characteristics in the combined cohort at different MELD and ANGPT2 threshold values. (A) Sensitivity/specificity plot showing the performance of ANGPT2 on day 1 (n=42 poor outcome, n=60 survivors). (B) Graph demonstrating accuracy across the range of ANGPT2 levels. The light grey represents the 95% CI. (C) Table summary of key performance values: sensitivity, specificity, and accuracy. MELD cutoff values were chosen from the literature and ANGPT2 values were selected as the values either optimized the sum of sensitivity and specificity or accuracy (ANGPT2 value of 10975 pg/ml resulted in both the highest accuracy and sensitivity/specificity at Day 1). (D) Sensitivity/specificity plot of ANGPT2 at Day 3 (n=38 poor outcome, n=53 survivors). (E) Accuracy plot. (F) Table of performance metrics.

**Figure 7:**
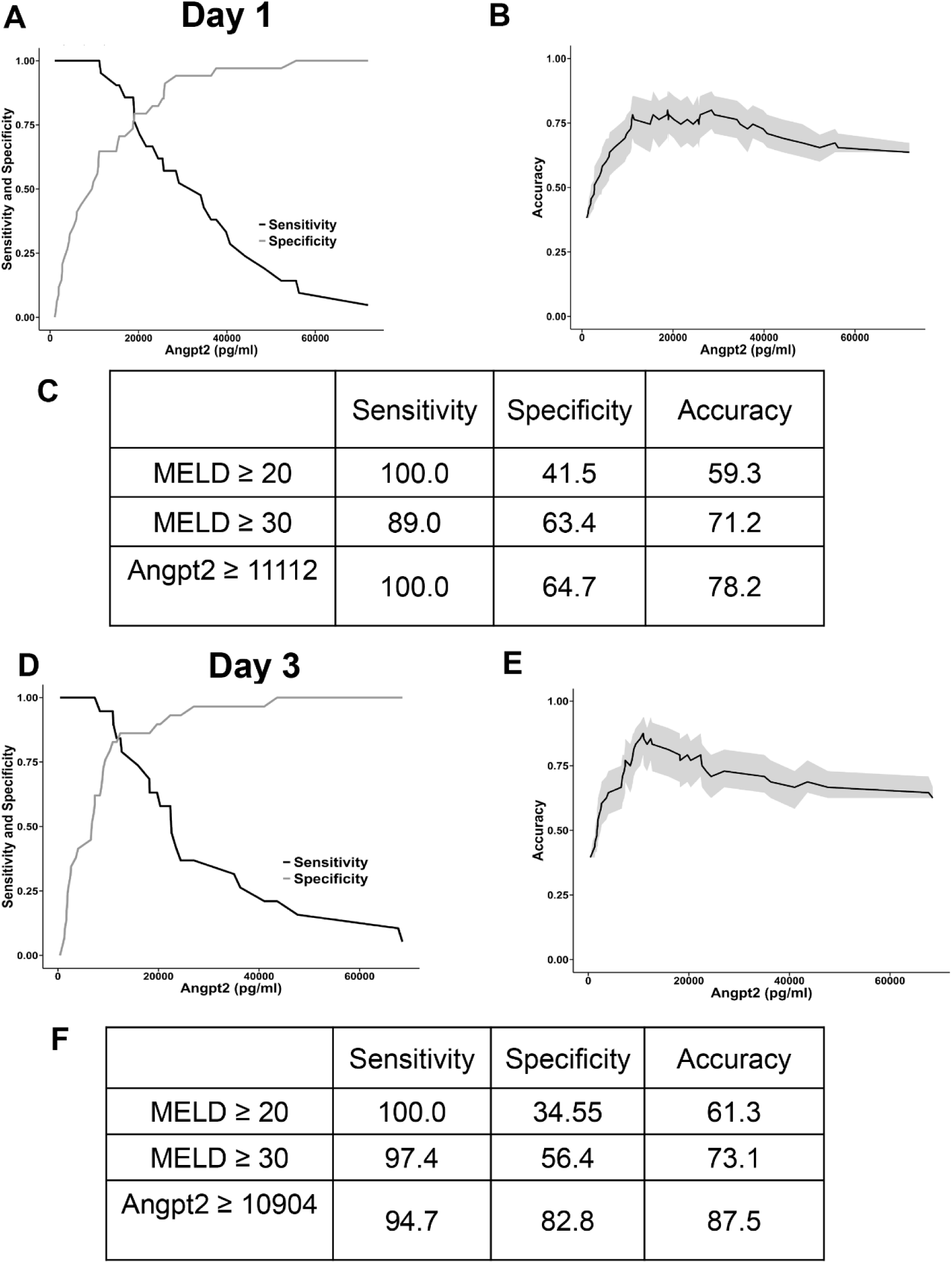
Performance characteristics in the subset cohort at different MELD and ANGPT2 threshold values. (A) Sensitivity/specificity plot showing the performance of ANGPT2 (n=21 poor outcome, n=34 survivors). (B) Accuracy plot demonstrating overall accuracy across the range of ANGPT2 levels. Light grey represents 95% CI. (C) Table summarizing key threshold values: sensitivity, specificity, and accuracy. MELD values were from on the literature and ANGPT2 values were selected as the values either optimized the sum of sensitivity and specificity or accuracy. (D) Sensitivity/specificity plot of ANGPT2 at Day 3 (n=19 poor outcome, n=29 survivors). (E) Accuracy plot. (F) Table of performance metrics.

## DISCUSSION

Our findings suggest that ANGPT2, an endothelial-specific factor associated with angiogenesis and inflammation, serves as an effective prognostic biomarker for APAP-induced ALF. Elevated levels of ANGPT2 in non-survivors, as observed in both the Phoenix (Fig 2) and ALFSG (Fig 3) cohorts, highlight its potential to differentiate between patients with poor outcomes and survivors. Importantly, ANGPT2 outperformed MELD, a commonly used clinical score, particularly when stratifying patients based on the onset of symptoms relative to clinical presentation based on AUC (0.83, 0.87 vs 0.80, 0.83, ANGPT2 vs MELD, Day 1 and Day 3, respectively, Fig 5). The performance metrics of ANGPT2, both alone and in combination with MELD, underscore its prognostic utility (Combination Model: 0.85, 0.90, Day 1 and Day 3, respectively, Fig 5). The stable threshold values for ANGPT2 further emphasize its potential as a reliable biomarker for guiding clinical decisions. The improved performance of ANGPT2 in early presenting patients suggests that it could be particularly useful for timely intervention in clinical settings.

In mouse models of APAP hepatotoxicity, there is an established temporal sequence of biological events: APAP bioactivation into the reactive metabolite, NAPQI (32), depletion of intracellular GSH (33), mitochondrial dysfunction (34), necrotic hepatocyte death, the release of DAMPs (35), initiation of a sterile inflammatory response (36) and ultimately, hepatocyte proliferation and recovery (37). Many old and new biomarkers can be tied to this sequence of events, such as proxies of hepatocyte injury like ALT/AST, LDH (16), DAMPs such as GLDH, mitochondrial DNA, nuclear DNA (11, 14), and HMGB1 (38), and proxies for hepatocyte regeneration such as AFP, osteopontin (15), and an early time-point miRNA signature (30).

Other biomarkers like CPS1 and mitochondrial DNA represent hepatocyte injury but also may specifically delineate a mitochondrial-associated injury (11, 12, 14). However, the development of APAP-induced ALF is poorly predicted by the magnitude of hepatocyte injury, for example, as reflected in transaminase levels, suggesting that other concurrent biological events in non-parenchymal cells may be a necessary requisite for poor outcome. Many of the known APAP-induced pathophysiological mechanisms occur in ECs, including GSH depletion, peroxynitrite formation and EC death (39, 40). Our evaluation of multiple human datasets revealed that *Angpt2* is highly expressed in ECs (Supp Fig 1), supporting that ANGPT2 in circulation may be a useful proxy measure for EC dysfunction.

ANGPT2 binds the Tie-2 receptor and has been shown to destabilize blood vessels, increase capillary leakiness and promote an endothelial response to inflammatory signaling (41).

Numerous investigations have considered ANGPT2 to be an endothelial biomarker in severe acute kidney injury (AKI) with sepsis (42,43) and linked ANGPT2 to organ failure and mortality (42,44,45). Interestingly, ANGPT2 can activate neutrophils, promoting adhesion onto ECs and influence platelet-activating factor (PAF) production (46,47). We previously reported a dose-dependent role for neutrophils in the APAP mouse model, whereby following a severe APAP overdose (600 mg/kg), neutrophils exacerbated liver injury, while after a moderate APAP overdose (300 mg/kg), neutrophils did not influence injury (48). The precise mechanistic underpinnings remain to be fully elucidated, however, neutrophils have been shown to promote hepatic platelet accumulation in mice treated with 600 mg/kg APAP (49), and platelet accumulation in the liver is associated with more severe injury (50). Moreover, neutrophil killing is critically dependent on cell adhesion through β2-integrins (CD18) (51), which can be induced on ECs exposed to ANGPT2 (52). Taken together, these investigations highlight a potential crosstalk initiated by ANGPT2 between ECs, neutrophils, and platelets that may coalesce to influence patient outcome.

In our recent work we identified the chemokine CXCL14 as a novel prognostic biomarker for poor outcome in APAP-induced ALF (17), linking the secretion of CXCL14 to the exacerbation of critical mechanistic events in the subset of APAP overdose patients with poor outcome (18). By applying a similar methodological framework, we have identified exacerbated endothelial stress as another crucial factor which can be linked to a proxy measure in the blood to predict patient outcome. We propose that by continuing to understand the mechanistic underpinnings beyond hepatocyte toxicity in APAP-induced ALF, further progress can be made to identify new biomarkers which can offer biological insight into the pathophysiology.

In conclusion, ANGPT2 is a promising prognostic biomarker for APAP-induced ALF, reflecting the exacerbated endothelial stress in patients with poor outcomes. Its superior performance compared to MELD, particularly in early presenting patients, highlights its potential for clinical application in improving patient prognosis and guiding therapeutic interventions. Further studies are warranted to establish its utility in diverse patient populations and clinical settings.

## Data Availability

All data produced in the present study are available upon reasonable request to the authors

## Conflict of interest

The authors declared no potential conflicts of interest with respect to the research, authorship, and/or publication of this article.

## Funding information

This work was funded in part by the National Institute of Diabetes and Digestive and Kidney Diseases (NIDDK) grants R01 DK102142 (H.J.) and DK125465 (A.R.), and National Institute of General Medicine (NIGMS) funded Liver Disease COBRE grants P20 GM103549 (H.J.) and P30 GM118247 (H.J.). D.S.U. was supported by an NIH Predoctoral Fellowship (F31 DK134197). The ALFSG is funded by the National Institutes of Health (NIH) grant (U01DK058369 to W.M.L.)

## Authors’ Contributions

David Umbaugh, MS (Conceptualization: Supporting; Formal analysis: Lead; Data acquisition: Supporting; Writing– original draft: Lead; Writing– review & editing: Supporting)

Nga T. Nguyen, PhD (Conceptualization: Supporting; Methodology and Data acquisition: Lead; Formal analysis: Supporting; Writing– review & editing: Supporting)

Steven C. Curry, MD (Patient recruitment: Lead; Writing– review & editing: Supporting) Jody A. Rule, PhD (Data acquisition; Supporting; Writing– review & editing: Supporting) William M Lee, MD (Patient recruitment and Funding acquisition: Lead; Writing– review & editing: Supporting)

Anup Ramachandran, PhD (Conceptualization: Supporting; Formal analysis: Supporting; Writing– review & editing: Supporting)

Hartmut Jaeschke, PhD (Conceptualization: Lead; Formal analysis: Supporting; Funding acquisition: Lead; Supervision: Lead; Writing– review & editing: Lead.

## Notes

### Competing Interest Statement

The authors have declared no competing interest.

### Author Declarations

The patient samples and data for both cohorts were acquired with informed consent and approved by the Institutional Review Boards of the Banner University Medical Center Phoenix, Phoenix, AZ, and the University of Texas Southwestern Medical Center, Dallas, TX in adherence to the 1975 Declaration of Helsinki.

