## Supplemental Material for "The endothelial growth factor Angiopoietin-2 is an accurate prognostic biomarker in patients with acetaminophen-induced acute liver failure"

### Supplemental Figure 1

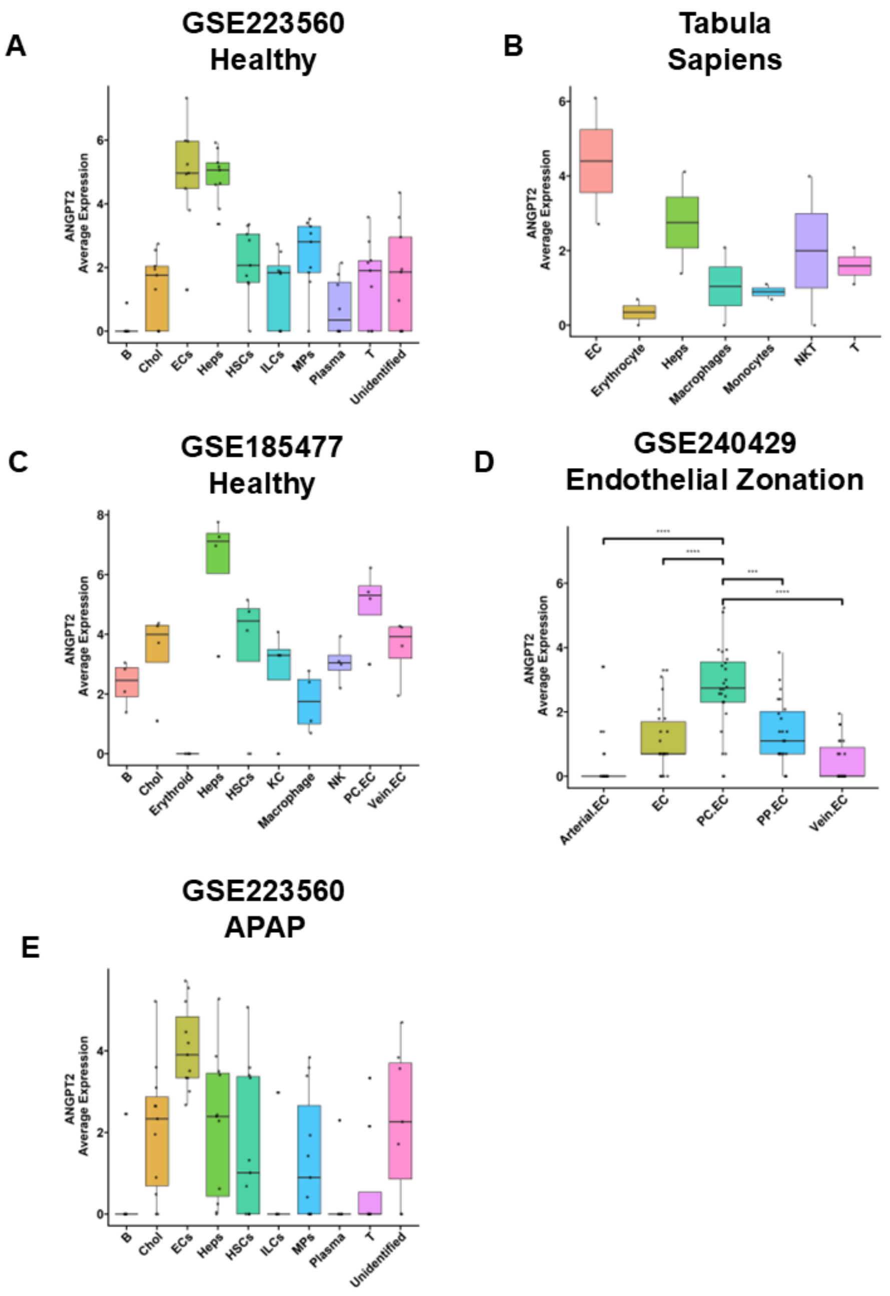


**Supplemental Figure 1: Evaluation of multiple human single-cell RNA sequencing datasets for Angpt2.** (A) Dataset was obtained from GEO at GSE223560 (n=9). (B-D) Datasets were obtained from the Chan Zuckerberg CELL by GENE Discover as processed and annotated scRNAseq data (B: n = 2, C: n=4, D: n=24). (E) Data was obtained at GSE223560 and filtered to include only the APAP samples (n=12). The y-axis is log-scale for visualization.

### Supplemental Figure 2

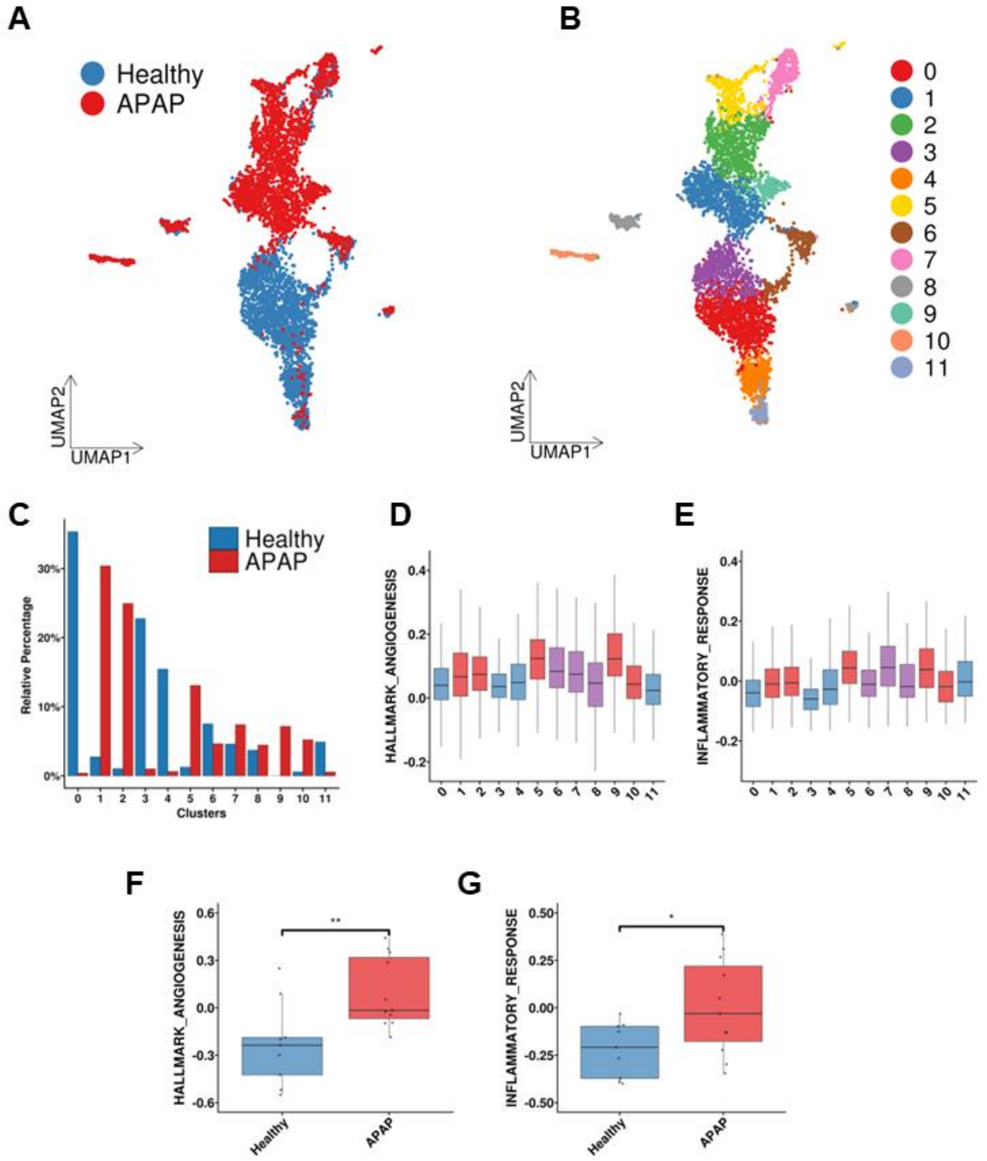


**Supplemental Figure 2**: **Analysis of human APAP single-nuclei RNA sequencing reveals the induction of angiogenesis and inflammatory pathways in endothelial cells.** (A) UMAP representation of human Single-nuclei RNA seq data GSE223560 consisting of liver explant from APAP overdose patients (APAP) or livers from patients who died of causes unrelated to liver (Healthy). (B) UMAP colored by cluster. (C) Bar graph showing the relative percentage of cells in each cluster. (D-E) Seurat's AddModuleScore was used to calculate the gene score from the HALLMARK_ANGIOGENESIS (D) and the WP_INFLAMMATORY_RESPONSE (E) for each cluster. The color represents the majority class: blue is healthy, red is APAP, and purple is mixed. (F-G) Boxplots demonstrating the gene score but at the individual sample level (healthy n= 9, APAP n = 12).

### Supplemental Figure 3

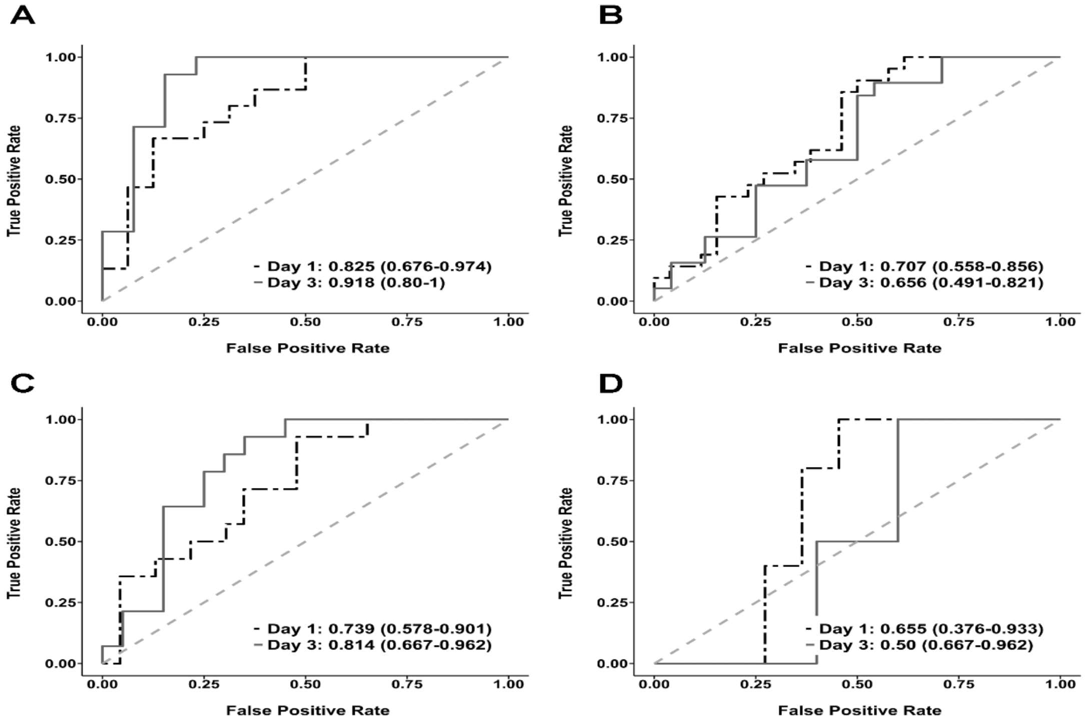


**Supplemental Figure 3: ALFSG cohort subset analysis demonstrates improved prognostic performance of ANGPT2.** (A) AUROC of ANGPT2 at Day 1 and Day 3 in ALFSG patients with reported onset of symptoms fewer than 3 days. (B) AUROC of ANGPT2 at Day 1 and Day 3 with reported onset of symptoms greater than 3 days. (C) AUROC of ANGPT2 at Day 1 and Day 3 in ALFSG cohort patients who presented to the clinic within 2 days or greater than 2 days (D) after the last consumption of APAP.
